# The impact of ethical implications intertwined with tuberculosis household contact investigation: a qualitative study

**DOI:** 10.1101/2024.06.27.24309538

**Authors:** LM Mlambo, M Milovanovic, CF Hanrahan, KW Motsomi, MT Morolo, MP Mohlamonyane, NW Albaugh, K Ahmed, NA Martinson, DW Dowdy, NS. West

**Author notes:** Corresponding Author: Nora S. West. **Parent Trial registration:** ClinicalTrials.gov: NCT04520113.

## Abstract

**Background:** Household contact investigation (HCI) is an effective and widely used approach to identify persons with tuberculosis (TB) disease and infection, globally. Despite widespread recommendations for the use of HCI, there remains poor understanding of the impact on and value of contact investigation for participants. Further, how HCI as a practice impacts psychosocial factors, including stigma and possible unintended disclosure of illness among persons with TB, their families, and communities, is largely unknown.

**Methods:** This exploratory qualitative study nested within a randomized trial (ClinicalTrials.gov: NCT04520113, 17 August 2020) was conducted in South Africa to understand the impacts of HCI on index patients living with TB and their household contact persons in two rural districts in the Limpopo province (Vhembe and Capricorn) and Soshanguve, a peri-urban township in Gauteng province. People with TB and household members of people with TB were recruited to participate in in-depth interviews and focus group discussions using semi-structured guides. We explored individual, interpersonal, and community-level perceptions of potential impacts of household contact investigation to elucidate their perceptions of HCI. Thematic analysis identified key themes.

**Results:** Twenty-four individual interviews and six focus group discussions (n=39 participants) were conducted. Participants viewed HCI as an effective approach to finding TB cases, helpful in educating households about TB symptoms and reducing barriers to health-related services. At the interpersonal level, HCI aided people with TB in safely disclosing their TB status to family members and facilitated family and social support for accountability. The introduction of HIV testing during HCI was reported by some participants as making household members slightly uncomfortable, decreasing interest in household members being tested for TB. HCI negatively impacted community-level TB and HIV-related stigma due to healthcare worker visibility at home.

**Conclusion:** Our data suggests varying impacts of HCI on people with TB, their families and interpersonal relationships, and communities, highlighting the importance of considering approaches that address concerns about community stigma and HIV testing to enhance acceptance of HCI.

## Background

The World Health Organization (WHO) has proposed an ambitious strategy to end the global TB epidemic by 2035 (1). To meet this goal, urgent attention is needed to factors that may influence the success of routine programmatic approaches, such as screening of household members of people diagnosed with TB.

Identifying people with TB and those at substantial risk of TB infection remains essential for global health and development (1, 2). Most people with TB are diagnosed passively by presentation at health facilities (3). However, persistent barriers contribute to clinic attendance: being asymptomatic (e.g., subclinical or early TB disease) (3), lack of knowledge about TB or TB symptoms (4, 5), financial costs (6), transport availability, and facility proximity (7, 8). A common approach to ensuring contact persons access TB screening and are linked to care is household contact investigation (HCI) (9). Though approaches may vary by setting, HCI typically involves healthcare workers visiting the home of a newly diagnosed person with TB and screening and/or testing household members for TB symptoms. However, HCI may cause potential ethical challenges. Though scant, there is some evidence that HCI may increase stigma, discrimination, and blame for people with TB; a better understanding of these challenges can help ensure that any potential harms are mitigated (10–12).

Previous studies conducted in South Africa have found that screening for TB in the home setting, though often associated with psychosocial and logistical challenges, may reduce barriers to testing other household members (13, 14). As HCI is increasingly prioritized in high-burden settings like South Africa (15–17), understanding the ethical implications associated with HCI is critical to ensuring that TB researchers and program planners alike address the less tangible benefits and potentially harmful impacts of HCI. Therefore, the goal of this study was to use qualitative methodology to explore how stigma, confidentiality, and disclosure manifest in the context of HCI and impact people diagnosed with TB, their household members, and interpersonal relationships within the household and community in rural and peri-urban settings in South Africa.

## Methods

### Study Setting and Design

We conducted an exploratory qualitative study in South Africa to understand the impacts of HCI, with a focus on ethical factors, among people living with TB and their household members. Focus group discussions (FGDs) and in-depth interviews (IDIs) were used to collect data on both individual and group-level perceptions. This study was nested within a larger randomized trial of two novel TB HCI intervention approaches: the Kharituwe trial. The two novel TB HCI interventions included 1) HCI on holidays in rural areas and 2) HCI on evenings and weekends in urban areas compared to the standard HCI (during working hours). The trial was conducted among people with active TB and their household members in 12 hospitals across Vhembe and Capricorn districts in Limpopo Province, plus 33 clinics and two hospitals in Soshanguve and surrounding areas (a mix of urban and peri-urban communities spanning the borders of Gauteng and North West provinces). Eligibility and recruitment procedures for the trial are described elsewhere (18).

### Participant Recruitment and Sample

Potential participants aged 18 years and older were purposively selected from among Kharituwe trial participants and approached to consider providing informed consent, which was obtained before being interviewed in either a focus group or an interview, with attention to ensuring a balance of recruitment between sites as well a balance of gender and age within the sample.

### Data Collection

From June 12^th^, 2021, to February 23^rd^, 2022, we conducted 24 IDIs among people living with TB (n=12) and people living in their households (n=12), plus six FGDs with people living with TB (n=15) and TB household contact persons (n=24). IDIs and FGDs were conducted in English, Tshivenda, Sepedi, Tsonga, or Setswana languages and facilitated by experienced qualitative interviewers and focus group leaders who received additional training on the study aims and data collection instruments. All IDIs and FGDs were conducted in a confidential space at either the local health facility or study offices.

For interviews, the study team developed a semi-structured interview guide to elicit individual-level perceptions and insights on the experiences with and ethics of HCI (Supplementary File 1). Interview guides were structured to explore the following domains in the context of HCI: TB/HIV disclosure preferences, perceived stigma/blame associated with testing for TB or having a new TB diagnosis, community stigma or blame resulting from HCI; right to privacy; the relative importance of household contact investigation when weighed against potential negative impacts; willingness to participate in household contact investigation; family and household dynamics; duty to others health within the household; and beliefs about TB/HIV confection. For focus groups, the study team developed a facilitator guide to elicit discussions on community-level perceptions and norms regarding the ethics of household contact investigation (Supplementary File 2). Focus group guides were structured to explore the following domains: methods for reducing TB-related stigma among people with TB and their families; belief as to whether contact investigation should be mandatory/optional/not performed; and relative stigma of TB and HIV and the risk of stigmatizing people for either TB or TB and HIV by offering HIV testing as part of HCI.

### Analysis

A senior researcher conducted thematic analysis, M.M., experienced in qualitative methodologies, and a junior researcher, L.M.M., with support from N.S.W. A list of initial codes was generated by (M.M) and reviewed by (N.S.W) based on the study aim and questions from the semi-structured interview and focus group discussion guides. New codes were developed during analysis in response to emerging themes. This strategy allowed for an inductive and deductive approach to coding and analysis. Two South African analysts (L.M.M. and M.M.) reviewed the codebook to ensure agreement before coding, and it was re-assessed throughout the coding process. Both analysts had knowledge about the study but were not part of the parent study or data collection process.

The codebook and transcripts were imported into ATLAS.ti© analytical software for coding. Coding was done by (L.M.M.) with regular discussion with the study team regarding data interpretation. Codes were connected and sorted into identified themes and summarised into a tabular format with participant quotes. Two analysts (L.M.M and M.M) reviewed the tables to refine themes further. The analysts practiced reflexivity throughout the analysis by discussing and acknowledging their position and experiences as South African TB researchers and their experiences with and perceptions of HCI. Findings were examined by data collection method (IDIs, FGDs) and location but are not differentiated as such here unless noted because of consistency across these categories.

Final themes were reviewed and discussed by the entire analysis team to provide a deeper understanding of the contribution of each theme to the overall research goals (19). A social-ecological model was used to situate the results and demonstrate the multilevel ethical factors intertwined with HCI for individuals, households, and communities (20, 21).

### Ethics statement

The study was approved by the Human Research Ethics Committee at the University of the Witwatersrand (Ethics reference: 1909111B), and by the Institutional Review Board at the Johns Hopkins Bloomberg School of Public Health (IRB reference: 11124). All participants provided written informed consent before enrolment.

## Results

IDI participants included ten men and 14 women with a median age of 32 (IQR 29,55) years. Focus group discussion participants were 20 men and 19 women with a median age of 43 (IQR 30,60) years. Participants included 27 people diagnosed with TB and 36 household members (participant demographics are presented in Table 1, see Appendix 1). Overall, people with TB and household members of people with TB described varied perceptions of the impact of HCI in the context of critical ethical considerations. Participants discussed the largely positive role of HCI in health service delivery and access and aiding in fear reduction by delivering information about TB and transmission prevention at the household level. By contrast, participants also noted the negative impact HCI has concerning the perceptions of concurrent TB and HIV testing as well as TB and HIV-related stigma. Figure 1 presents key themes within the social-ecological model.

**Figure 1.**
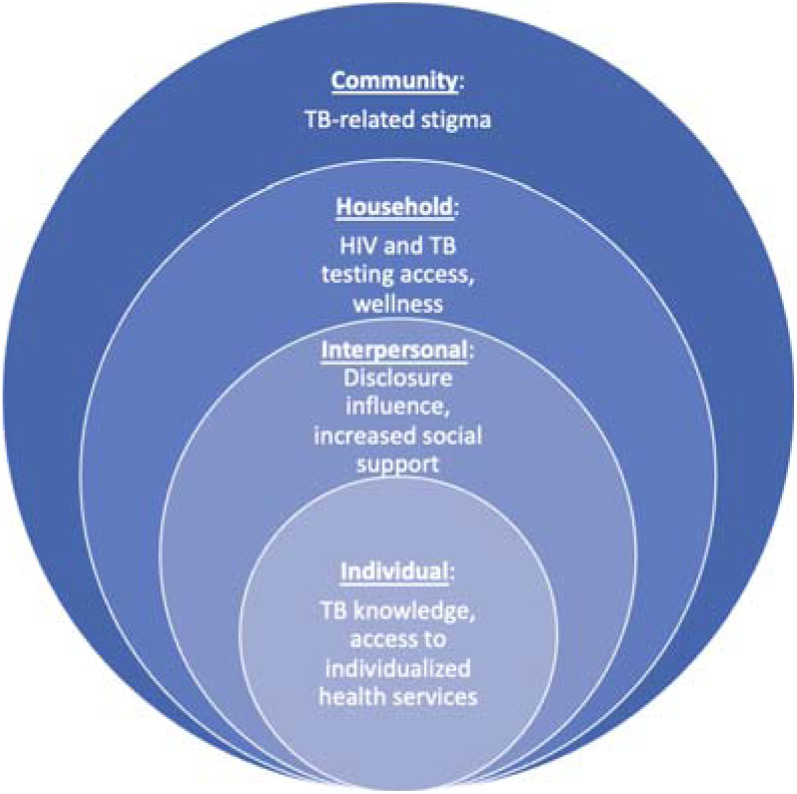
Key themes situated in the social-ecological model. Thematic analysis was used to derive key themes from the FGDs (n=6) and the IDIs (n=24). The key themes are presented within the social-ecological model to understand the range of ethical factors intertwined with the implementation of HCI within the individual, community, household, and interpersonal levels. HCI had a positive impact on the individual and interpersonal level, providing education and access to health services as well as aided disclosure of TB status and social support. Within household and community levels, HCI decreased interest in testing for HIV and increased TB and HIV-related stigma.

### HCI as a means of reducing barriers for individuals accessing services

According to participants, HCI was viewed positively, playing a pivotal role in ensuring that people with TB and their household members receive timely access to care and TB-related services. People with TB and their household members described HCI as cost-saving, reassuring, and less time-consuming than seeking TB screening through the clinic.

> *“I think it’s important that they often come and check on us because some people don’t like going to the clinic; from what I have seen, it’s difficult because we almost lost a person… I think it’s good because at home, we will be thinking that he [household member with a TB diagnosis] is the one who is sick and we are not sick. So he is the one who needs to get help, but when they came at home they were also able to assist us and we got assistance. Because they are also saving us transport to go to the clinic for all of us to go and get tested.”* – Household member

HCI was considered impactful by many participants because it reached key groups like older adults or individuals who might be sick but for various reasons (e.g., unaware of TB symptoms, perceived their illness as minor, or feared being seen by the community at the clinic) were hesitant to seek care. Therefore, HCI was perceived as one of the most beneficial programs within the health system to reduce barriers to accessing TB-related screening and health services.

### Increased individual TB knowledge and reduced fear

Both people with TB and household member participants noted that at the level of the individual, HCI provided direct knowledge about TB and taught household members how to live with someone who has TB, which could, in turn, positively impact interpersonal relationships and potentially minimize within-household discrimination. A participant described how HCI provided reassurances to people with TB and their household members who had concerns about transmitting or contracting TB:

> *“The house visits also motivate your family… Because you are bringing education to them, so that they know that this person is our patient, and what you need to know is that you must live with this person like this, and like that… And you as well, as they have explained to you that you need to do this and that so that you don’t infect others…”* - Person with TB

### Interpersonal relationships: duty, responsibility, disclosure, and support

Many participants, both people with TB and household members, described the importance of family health and how HCI influenced communication and actions among household members. First, some participants described the knowledge that HCI visits would be conducted as a catalyst for the disclosure of TB status to the family. However, others reported disclosing their status to their family members regardless of whether a HCI visit was made. Importantly, disclosing one’s TB status was believed to undoubtedly alleviate the stress of suffering alone because “*a secret sometimes kills.”* Second, many participants emphasized that knowing HCI would take place helped people with TB consider household safety and their role in decreasing the likelihood of TB transmission to others.

Third, participants strongly expressed that the person with TB had a responsibility to disclose to others and play a role in the prevention of TB and screening for household members, which HCI facilitated:

> *“…You either protect this household people that live with a person with TB too. As they say, prevention is better than cure. If they don’t have TB, then they have to know that they don’t have TB. And then if they have, they can be tested for that too.”*-Person with TB

Despite the perceived beneficial role that HCI played in facilitating disclosure of TB status, some participants noted that disclosure could also result in discrimination within the household. Most participants viewed a person with TB rejecting HCI for their household as the result of either unawareness of the infectiousness of TB or associated with fear of negative household or community perceptions. For many participants, HCI as an approach was described as something that strongly enhanced a sense of responsibility for the household and personal health.

Some people with TB felt that HCI could often reduce household members’ concerns about TB infection by providing the necessary health information, thus facilitating a cohesive and supportive space in which to care for persons living with TB.

> *“We are the ones that benefit. Like you are teaching us about TB. They are also telling us [about TB] at the clinics as well, but they only telling us half [of the information], right? Like with you guys, you encourage [educate] us, on how to live, what to eat…How to live with the people we live with in the household.”*-Household member

Participants felt that the motivation to disclose TB status and subsequent counseling received from HCI helped increase the level of family support offered to people with TB by empowering household members to hold people with TB accountable for adhering to treatment, assist with the collection of medication, and provide support through the provision of meals.

### Household uptake of HIV Testing during HCI

HIV testing is a standard service often offered as part of the care package during a HCI visit. Participant perceptions of the HCI approach were intertwined with feelings about HIV testing. Both people with TB and household members described how HIV testing being linked to HCI could negatively impact a household’s decision to participate in HCI:

> *“…TB currently; when you go test for it, they automatically test your HIV status… I think a person who is not ready for that. If they haven’t fallen sick, hasn’t had any symptoms… that is what could make them say they aren’t ready for [HIV testing]…So, you find that even if they want to check/test for TB only, they will say that they also going to test me for [HIV]-… So, I am not ready…”* – Person with TB

Participants felt that household members who would be less comfortable with the HCI approach also feared *“knowing the truth”* regarding their HIV status. Despite the noted challenge concerning the impact of offering HIV testing on TB screening/testing uptake, most participants acknowledged the importance of testing for HIV. Further, many participants also described that HIV testing as part of HCI may offer privacy, discretion, and convenience as described by a household member of a person with TB:

> *“When they say ‘can we also check you on this’ they are not only helping themselves but they are also helping you. That’s why you have to agree so that you can know where you stand, yes.”* – Household member

### The Role of Community Stigma

Most participants considered HCI an important approach for finding new cases of TB and recognized the importance of identifying people with TB in their households or communities; however, nearly all participants raised concerns about community perception and potential stigma. Descriptions of stigma primarily focused on the community, as opposed to stigmatization within the household. Overall, HCI was perceived to contribute to or exacerbate illness-related stigma because healthcare workers visiting households in marked cars and typically wearing uniforms were viewed as demonstrating to neighbors or other community members that someone within the household was sick. Many participants described being seen by others with healthcare workers visiting the household as potentially distressing because this might change a person’s ‘social identity’, labeling them as someone with an illness.

> *“…like when they come, we know that obviously, it’s those people wearing like obviously maybe those blue things or whatever… Yes, that uniform of theirs… They can see that “oh it means that in there, like when they [the healthcare workers] go there each and every day–… it means there is someone who is ill in there [at that particular house]”* … So, people are ashamed because of that thing…” - Household member

Within discussions of stigma, participants described two distinct but overlapping types of potential stigma: 1) HIV-related stigma, noting that people with TB were sometimes believed to have HIV because of known high co-occurrence and overlapping symptoms such as loss of weight and coughing, and 2) TB-related stigma. Several participants described how their communities were uninformed about TB, including the medical journey from TB diagnosis to treatment and the likelihood of being cured. Participants strongly emphasized that HCI could play a role in educating people about TB and HIV.

> *“…Just judging…So automatically, when they see you coughing or see that you have lost weight or you have TB, they say ‘oh well she/he says it is TB, but she/he knows what it goes together with [HIV]’…it depends on people’s understanding about that thing [TB and HIV]…A person who doesn’t understand, who has little knowledge, is going to combine them…But one who has full knowledge and a lot, won’t combine them…The thing about when we get to the homes, we have to educate people…”* – Person with TB

Overall, participants noted that the stigma driven by HCI may isolate people and households with TB because fear of TB and HIV-related stigma is pervasive in communities.

## Discussion

The findings from this qualitative study demonstrate that HCI, a recommended approach to identifying people with TB, is perceived as having largely positive impacts at the household level but potential negative implications concerning community and HIV stigma. HCI fosters social support, reduces barriers to screening and healthcare engagement, and may facilitate the disclosure of TB diagnosis. However, the HCI approach is perceived as a potential driver of health-related stigma within the community. Overall, HCI as a programmatic approach has impacts beyond identifying and screening individuals for TB, spanning the interpersonal, household, and community levels.

Governments must deliver TB services, fulfilling the human right to healthcare (11). However, traditional or “passive” identification of people with TB (e.g., relying on individuals to go to the clinic for screening or care) presents challenges, including access to the health facility for timely identification of TB symptoms (3). A study in South Africa on healthcare access found that most individuals, particularly those living in low-income communities, postponed seeking healthcare due to distance from facilities and travel costs (7). Many participants in our study felt HCI had an essential role for the individual because it reduced noted structural and economic barriers to TB screening. Both persons with TB and household members highlighted how HCI, compared to clinic-based care, helps avoid long queues (5) and alleviates travel costs, (7, 8) thus improving the timely screening of TB and supporting the use of HCI in high-burden TB settings.

In our study, participants emphasized the utility of HCI in educating households, including household members and people with TB, on TB symptoms, treatment, and transmission. A study conducted in Free State, South Africa, found that educating people with TB about the disease positively influenced treatment adherence (4). Additionally, household TB knowledge is crucial for understanding TB infectivity and supporting prevention behavior (15). In Uganda, TB knowledge helped people with TB understand and make informed decisions, including protecting themselves and their close contacts from TB transmission (22). People with TB have reported experiencing stigma and withheld support within the household (23); in our study, participants described how HCI facilitated the sharing of TB diagnosis with close contacts despite the potential stigma. HCI also provides both education and reassurance to household members and people with TB, which may help mitigate household-level stigma (6, 23). In this case, decision-makers must balance the need to educate and counsel persons with TB and reduce barriers to testing their close contacts against the potential unintended adverse effects of increasing community-level stigma (11).

Stigma continues to obstruct the fight against TB, including the implementation of interventions such as HCI (24). For example, studies conducted in South Africa assessing the level of vulnerability and stigma experienced by people with TB found that TB interventions (both passive case finding and contact tracing) were often hampered because of factors such as negative community perceptions, the anticipated HIV-TB stigma and the association of TB with dirt (24, 25). Similar to other studies conducted in South Africa (3), our findings support evidence that people with TB and household members have mixed feelings about healthcare workers conducting household visits; specific concerns included the use of marked cars, uniforms that identify visiting healthcare workers, and neighbor perceptions that there is a sick person in the household. Ultimately, screening programs can unintentionally disclose the identity of people with TB (26), and while participants in our study described the benefits of HCI within their household as significant, concerns about community stigma – both HIV and TB-related - tempered enthusiasm for the approach. TB and HIV stigma ultimately remain significant yet are under-addressed problems in TB programming. Given that HCI programs potentially play a direct role in increasing stigma in communities for people with and affected by TB, stigma-mitigating interventions are crucial to prevent shame and discrimination. Such interventions can include educating and providing psychosocial interventions (e.g., psychosocial support groups or TB clubs) targeted towards communities, healthcare workers, and people living with TB; training healthcare workers on sensitivity and stigmatization; and considering policy interventions to measure and reduce TB stigma (21, 24, 27).

In line with other studies in high TB and HIV burden settings, our findings demonstrate that stigma among people with TB may be driven by the assumption that all individuals with TB are also living with HIV (24, 25, 28). In South Africa, efforts to integrate TB and HIV screening have led to the use of cost-effective methods like HCI to identify more individuals with both TB and HIV, reduce transmission, and improve clinical outcomes (29, 30). While this is a critical approach given that an estimated 71% of people diagnosed with TB in South Africa also have HIV (31), HIV-associated stigma may negatively impact the uptake of TB services (12, 32). A study conducted in Eastern Cape, South Africa, reported that people with TB were often assumed to be living with HIV, which impacted TB treatment adherence and care by discouraging people from going to the clinic for care and complete treatment (28). Importantly, we found that the association between TB and HIV was a factor that could influence a household’s decision to accept or reject HCI because of the provision of HIV testing. While integrated screening intends to reduce the burden of both TB and HIV, participants described significant HIV stigma concerns and the pressure driven by HCI to test for HIV. These findings demonstrate that, while offering HIV testing with HCI in high co-burden settings is a reasonable approach from a disease prevention standpoint (33), the impact of this practice on HCI uptake remains poorly understood. Filling this knowledge gap will allow screening programs to make informed decisions about the provision of HIV testing as part of HCI and ensure HCI programs are people-centered.

## Limitations

This study has limitations. First, South Africa is among the top 30 high-burden HIV/TB countries; thus, these findings may not generalize to settings with high TB burden but low HIV prevalence. Second, participants were sampled from a parent contact investigation study, which may have influenced opinions of the HCI approach compared to individuals with no exposure to HCI. However, as HCI is a commonly used approach in South Africa, study participants’ views may not differ significantly from those of individuals not engaged in research. Finally, we did not find differences across the geographical areas (urban vs. rural) as expected, which may be the product of implementing a set of standardized questions that were not necessarily site-specific. However, we explored data across participant types and data collection methods (FGDs and IDIs), strengthening the rigor of our findings.

## Conclusion

HCI as an approach has varying impacts on people with TB, their families, interpersonal relationships, and communities. This qualitative study suggests that HCI is broadly acceptable with perceived positive impacts on household health and TB knowledge, TB status disclosure, and social support for people with TB. However, the intersection of HCI and TB and HIV-related stigma is of critical importance to individuals impacted by TB and has been largely unaddressed in HCI implementation to date. The provision of HIV testing, in particular, may have negative consequences on the uptake of TB screening and needs to be better understood. If HCI is to be scaled up, more significant efforts should be made to ensure concerns about community stigma and HIV testing are addressed.

## Data Availability

Data cannot be shared beyond individual quotations in the manuscript because participants did not give consent for the publication of their responses in full, or access to their data to those outside of the research study will have access to participants' data. Requests for data for researchers who meet the criteria for access to confidential data will be considered by the governing IRB on a case by case basis.

## Lists of abbreviation

HCI: Household Contact Investigation
TB: Tuberculosis
HIV: Human Immunodeficiency Virus
WHO: World Health Organization

## Declarations

### Ethics approval and consent to participate

The study was approved by the Human Research Ethics Committee at the University of the Witwatersrand (Ethics reference: 1909111B) by the Institutional Review Board at the Johns Hopkins Bloomberg School of Public Health and conducted in compliance with the Declaration of Helsinki. All participants signed an informed consent form before being interviewed for this study.

### Consent for publication

Not applicable.

### Authors’ contributions

LMM: Formal analysis, data curation, writing – original draft and editing; MM: Formal analysis, writing – original draft and editing; CH: Investigation, conceptualization, writing – review and editing; KWM: Study administration, data curation, writing – review and editing; MTM: Study administration, data curation, writing – review and editing; MPM: Study administration, data curation, writing – review and editing; NWA: Study administration, data curation, writing – review and editing; KA: Conceptualization, supervision, writing – review and editing; NM: Conceptualization, supervision, study administration, writing – review and editing; DWD: Investigation, conceptualization, funding acquisition, methodology, writing - review & editing, NSW: Methodology, study administration, formal analysis, data curation, writing – original draft and editing.

## Acknowledgments

The authors thank the participants for their time and valuable insights. We thank C Ramsammy and J Coetzee for their help refining themes.

### Appendix 1. Participant demographics

| DISTRICT | PARTICIPANT TYPE | AGE RANGE<br>(years) | GENDER | HIV STATUS |
| --- | --- | --- | --- | --- |
| **In-depth Interviews** |  |  |  |  |
| Soshanguve | Person with TB | 40-59 | M | Negative |
| Soshanguve | Person with TB | 25-39 | F | Positive |
| Soshanguve | Person with TB | 40-59 | F | Positive |
| Soshanguve | Person with TB | 60+ | M | Positive |
| Vhembe | Person with TB | 18-24 | M | Negative |
| Vhembe | Person with TB | 25-39 | F | Positive |
| Vhembe | Person with TB | 40-59 | F | Positive |
| Vhembe | Person with TB | 25-39 | M | Negative |
| Capricorn | Person with TB | 60+ | F | Negative |
| Capricorn | Person with TB | 25-39 | M | Negative |
| Capricorn | Person with TB | 25-39 | F | Negative |
| Capricorn | Person with TB | 25-39 | M | Negative |
| Soshanguve | Household member | 25-39 | F | Positive |
| Soshanguve | Household member | 25-39 | F | Positive |
| Soshanguve | Household member | 60+ | M | Positive |
| Soshanguve | Household member | 25-39 | M | Positive |
| Vhembe | Household member | 60+ | F | Unknown |
| Vhembe | Household member | 40-59 | F | Negative |
| Vhembe | Household member | 25-39 | F | Negative |
| Vhembe | Household member | 25-39 | F | Positive |
| Capricorn | Household member | 40-59 | F | Negative |
| Capricorn | Household member | 25-39 | F | Negative |
| Capricorn | Household member | 60+ | M | Unknown |
| Capricorn | Household member | 18-24 | M | Negative |
| **Focus Group Discussions** |  |  |  |  |
| Soshanguve | Person with TB | 60+ | F | Positive |
| Soshanguve | Person with TB | 40-59 | M | Positive |
| Soshanguve | Person with TB | 60+ | M | Positive |
| Soshanguve | Person with TB | 40-59 | M | Positive |
| Soshanguve | Person with TB | 25-39 | M | Negative |
| Soshanguve | Person with TB | 60+ | M | Positive |
| Vhembe | Person with TB | 25-39 | M | Positive |
| Vhembe | Person with TB | 40-59 | M | Negative |
| Vhembe | Person with TB | 40-59 | M | Positive |
| Vhembe | Person with TB | 25-39 | F | Unknown |
| Capricorn | Person with TB | 40-59 | F | Positive |
| Capricorn | Person with TB | 40-59 | M | Negative |
| Capricorn | Person with TB | 25-39 | M | Negative |
| Capricorn | Person with TB | 25-39 | M | Negative |
| Capricorn | Person with TB | 25-39 | M | Negative |
| Soshanguve | Household member | 25-39 | F | Positive |
| Soshanguve | Household member | 40-59 | M | Positive |
| Soshanguve | Household member | 40-59 | F | Positive |
| Soshanguve | Household member | 40-59 | F | Positive |
| Soshanguve | Household member | 60+ | F | Positive |
| Soshanguve | Household member | 40-59 | F | Positive |
| Soshanguve | Household member | 60+ | F | Positive |
| Vhembe | Household member | 25-39 | M | Negative |
| Vhembe | Household member | 60+ | M | Unknown |
| Vhembe | Household member | 25-39 | F | Unknown |
| Vhembe | Household member | 25-39 | M | Unknown |
| Vhembe | Household member | 18-24 | F | Positive |
| Vhembe | Household member | 60+ | M | Positive |
| Vhembe | Household member | 60+ | F | Unknown |
| Vhembe | Household member | 18-24 | F | Negative |
| Vhembe | Household member | 25-39 | F | Positive |
| Capricorn | Household member | 40-59 | M | Positive |
| Capricorn | Household member | 60+ | F | Positive |
| Capricorn | Household member | 60+ | M | Positive |
| Capricorn | Household member | 40-59 | F | Positive |
| Capricorn | Household member | 25-39 | F | Positive |
| Capricorn | Household member | 25-39 | F | Negative |
| Capricorn | Household member | 25-39 | M | Negative |
| Capricorn | Household member | 18-24 | F | Positive |

## Supplementary File 1 Ethics of household contact investigation in South Africa: interview guide Kharituwe Bioethics Supplement

**In-depth Interview Guide Version 1.0/ 13 October 2020**

### Introduction

*Thank you for being here today. I want to remind you before we start talking, there are no right or wrong answers. I am here to learn from you and hear your ideas and suggestions*.

### Open-ended questions

*As you know, today we are going to speak about tuberculosis (TB), which is an infectious disease that mainly affects the lungs. Often people with TB have a cough, fever or weight loss – sometimes they may have TB for months or even years without knowing it. TB is spread from one person to another through the air. Because people often spend time with their household members, if one household member has TB and is coughing, other household members may also get TB. There is a cure for TB, and testing and treatment are free at the clinic*.

*One of the ways that is often used to find more people with TB and tell them about their treatment options is for health workers to visit the household of a person with a recent diagnosis of TB and check the other people who live in the household for TB. This may involve asking people if they have any of the symptoms of TB and taking a sputum sample from people who have TB symptoms. This way of finding new cases of TB is called household contact investigation. Often household members are also asked if they would like to be tested for HIV at these household visits*.

*Let’s start with getting some of your thoughts about household visits to check people for TB*.

1. I’d like to first start by asking what you think of health workers visit to the households of people who have TB to check other household members for TB?

Probe: Do you think it is important for health workers to visit the households of people who have TB to check other household members for TB?

Probe: Please explain to me whether you think visiting the households of people who have TB to check household members for TB is a good or bad way to find new cases of TB?

Probe: What would you propose as a different way to finding new cases of TB other than healthcare workers visiting homes to check household members for TB?

Probe: What are the benefits of visiting the households of people who have TB to check household members for TB?

Probe: What are some things that might not be good about visiting the households of people who have TB to check household members for TB?

Probe: If health workers do not visit the households of people who have TB to check their household members, what might happen?

2. [For index cases only]: Can you tell me how you felt or would feel about having someone come to your house to check other household members for TB?
[For contacts only]: If or when someone in your household was diagnosed with TB, can you tell me how you felt or would feel about having someone come to your house to check you and other household members for TB?

Probe: Do you think everyone in your household would feel comfortable having someone come to the house to check household members for TB? Why or why not? Which people would or would not feel comfortable with this, and why?

Probe: Please describe any concerns you would have about having someone come to the house to check household members for TB?

Probe: What are the reasons that people may or may not want to be checked for TB in the household?

3. I would like to hear your thoughts about, when a health worker comes to the household of someone recently diagnosed with TB to check other household members for TB, how that health worker should explain the reason for the visit. What do you think the health worker should say to the household members about the reason for their visit, and why?

Probe: Please tell me about the good or the bad things about the health worker saying the reason for the visit is because someone in the household has been diagnosed with TB and invited us to come.

Probe: Please tell me about the good or the bad things about the health worker saying the reason for the visit is because they are trying to test for TB in selected households in this community.

4. [Index case only]: Did you tell your household members that you have TB?? Why or why not?

Probe: (If yes to Question 4) Please tell me about your experience telling your household members that you were diagnosed with TB.

Probe [index case only]: Please describe anything that worried you about telling your household members that you had TB? Were there any household members who you decided not to tell? Why did you not want to tell these people?

Probe [index case only, if they did disclose]: Please describe how household members reacted when you told them about your TB diagnosis.

> [Contact only]: Were you told by anyone that a member of your household was diagnosed with TB? [If yes,] please tell me about your experience finding out that your household member had TB. [If no,] if someone in your household was diagnosed with TB, would you want to know – and if so, how would you like to be told? Please explain why.

Probe [contact only]: How did or would you feel when you found out or if you were to find out your household member had TB?

Probe [contact only]: How did or would you react when you found out or if you were to find out your household member had TB?

Probe [contact only]: Please describe any worries or concerns you had or would have when you found out or if you were to find out your household member had TB?

5. [Index case only]: Please tell me about the reasons for your decision to tell your household members that you had been diagnosed with TB?

Probe: Please describe to me if you had in mind the health of your household members when thinking about whether to tell them about your TB diagnosis.

Probe: Please tell me about anything that made you not want to tell your household members about your TB diagnosis.

Probe: Please tell me about how knowing that health care workers might come to your house influenced your decision of whether or not to tell your household members about your TB status.

6. [Index case only]: I would like to know more about your experience after you were diagnosed with TB. In general, how do you feel you are treated in your community because of your TB diagnosis?

Probe: Do your family members know about your diagnosis? How are you (would you be) treated by your family after learning of this diagnosis?

Probe: Do your close neighbors know about your diagnosis? How are you (would you be) treated by neighbors after learning of this diagnosis?

Probe: Do your close friends know about your diagnosis? How are you (would you be) treated by friends after learning of this diagnosis?

7. Please tell me about how you think having someone come to the house to check household members for TB might impact how you are treated in your community?

Probe: If people in your community notice someone is coming to the house to check for TB, please describe whether they might treat you differently?

Probe: Do you think their reaction might be different if they were visited by people in plain clothes and unmarked cars versus in uniforms and marked cars?

Probe: How do people in this community think about someone’s TB status in relationship to their HIV status? If they know that someone has been diagnosed with TB, will they assume that person has HIV as well?

Probe: Please describe how you think people in this community feel about someone who has TB?

8. Do you think people in this community would discuss their TB diagnosis with others? Why or why not?

Probe: What are the reasons someone might not want to tell others in the community that they have been diagnosed with TB?

*For the final part of our discussion, I am going to ask you about different timings of household contact investigation. Each involves having a health worker visit the household, but the timing of these visits are different in the three types. As I mentioned earlier, when I say household contact investigation I mean when health workers visit the household of a person with a recent diagnosis of TB and check the other people who live in the household for TB, and usually offer HIV testing as well. We are interested in your thoughts*.

*The first time that health workers could come and visit households after someone was diagnosed with TB is during normal business hours (9:00-17:00). This is what is currently done in places where TB contact investigation is done. A different time when health workers could visit instead would be in the evenings and on the weekends, when some people might be more likely to be home. And another possibility for timing, instead, is to visit on the holidays like the festive season or Easter. This holiday approach might have the benefit of having large families home during those times, but would also potentially involve delays (for example, if someone were diagnosed with TB in July and couldn’t be visited until December). Do you have any questions about these types of household visits before we go further?*

*Now let’s talk in more detail about each of these types of household visits…*

Note: Ask the following questions for each type of household visit (holiday, off-peak, and routine). Remind of participant of the definition for each type of household visit.

9. What do think about this type of visit to households of people who have TB to check other household members for TB?

Probe: What are the benefits of this type of visit to households of people who have TB to check other household members for TB?

Probe: What are the things that might not be good about this type of visit to households of people who have TB to check other household members for TB?

Probe: Please describe anything that would make you uncomfortable about this type of visit to households of people who have TB to check other household members for TB.

Probe: Please describe anything that would be good about this type of visit to households of people who have TB to check other household members for TB.

10. What do you think about offering HIV testing to others in the household when visiting the household during this type of visit to check for TB?

Probe: Please describe for me any problems there might be if health workers offer HIV testing at the same time as they come to check household members for TB.

Probe: Please tell me about anything that you think is good about having HIV testing offered to household members at the same time they are being checked for TB.

11. Thinking about all three types of household visits – holiday, off-peak, and routine – which approach do you think is best, and why?

Probe: Tell me about your choice…what would make that the best type over the others?

> *Thank you for your time…this information has been very valuable.*

## Supplementary File 2 Ethics of household contact investigation in South Africa: focus group discussion guide Kharituwe Bioethics Supplement

**Focus Group Discussion Guide Version 1.0/ 13 October 2020**

### Introduction

*Thank you all for being here today. I want to remind everyone before we start talking that there are no right or wrong answers. We are here to learn from you and hear your ideas and suggestions. We want all of you to tell us what you think and learn from you and your experiences*.

*―will lead the discussion today by asking a question to the whole group and call on people to get their thoughts. We want to hear from everyone. ― will be taking notes while we talk*.

*Does anyone have any questions before we start?*

### Open-ended questions

*One of the ways that is often used to find more people with TB and let them know about it is for health workers to visit the household of a person with a recent diagnosis of TB and check the other people who live in the household for TB. This may involve asking people if they have any of the symptoms of TB and taking a sputum sample from people who have TB symptoms. This way of finding new cases of TB is called household contact investigation. Often household members are also asked if they would like to be tested for HIV at these household visits*.

*Let’s start with getting some of your thoughts about household visits to check people for TB*.

1. As members of this community, please tell me what you think about having health workers visit the households of people who have TB?

Probe: Do you think it is important to have health workers visit the households of people who have TB? Why or why not?

Probe: Tell me about why you think this might be good for the households who are visited?

Probe: Tell me about why you think this might be good for the community?

Probe: How do you think having a health worker visit the household would make people feel?

Probe: Do you think people might feel pressured into doing something they don’t want to do? Why?

2. Thinking about all the benefits and negatives, please tell me about if you think that visiting households of people who are diagnosed with TB is something that should be done?

Probe: Do you think it’s something that clinics should focus on doing? Why or why not?

3. *I am going to present a scenario and I would like you all to tell me your thoughts about it:* Please tell me what you think about a scenario where there is a household and the people who stay there want to be checked for TB, but the person in the household who has been diagnosed with TB decides they do not want the health workers to visit the household to check everyone for TB.

Probe: What are the positive and negatives of someone with TB deciding that their household cannot be visited and other household members checked for TB?

Probe: In your opinion, is it fair or unfair for the person with TB to make this decision, and why?

4. *I am going to present another scenario and I would like you all to tell me your thoughts about it:* Please tell me what you think about a scenario where there is someone in the household who has been diagnosed with TB, and only some of the family members want to be checked for TB, but others do not want to?

Probe: Should the healthcare workers who are visiting ask people if they want to get tested for TB or tell them that they should get tested?

Probe: Please tell me your thoughts on the responsibility of household members who have had contact with the person who has been diagnosed with TB to get checked?

5. How does your community view people from outside of the community coming to someone’s house?

Probe: How do people in the community react or act when people from outside the community come to someone’s house?

6. Please tell me about how people in the community treat or act towards someone who they know has been diagnosed with TB.

Probe: Describe for me how people in the community treat or act towards someone they know has TB compared to someone who does not.

Probe: Describe for me how people in the community act towards household members of someone who has TB.

7. Do you agree or disagree that people in this community make assumptions about the HIV status of someone who has been diagnosed with TB or the HIV status of the household where someone has TB? Please explain.

Probe: What do people in the community assume about someone’s HIV status if they are diagnosed with TB, and why?

8. As members of this community, what do you think about offering HIV tests to all household members, when coming to visit a household for TB testing because a household member has TB?

Probe: What are the things that are good about offering HIV tests to all household members?

Probe: What are the bad things or negatives about offering HIV tests to all household members?

9. Please tell me your thoughts on how household visits to check for TB among household members could be made less shameful to patients and household members?

Probe: Tell me about things that the person or people visiting the home could do to not cause households to be stigmatized by individuals or the community?

*The first time that health workers could come and visit households after someone was diagnosed with TB is during normal business hours (9:00-17:00). This is what is currently done in places where TB contact investigation is done. The second time that health workers could visit would be in the evenings and on the weekends, when some people might be more likely to be home. The third time is on the holidays like the festive season or Easter. This approach might have the benefit of having large families home during those times, but would also potentially involve delays (for example, if someone were diagnosed with TB in July and couldn’t be visited until December). Do you have any questions about these types of household visits before we go further?*

Note: Make use of a flip chart to note the positives and negatives of each strategy. Reference back to the flip chart to remind people of each type of contact investigation for questions asking them to compare all three.

*Now let’s talk in more detail about each of these different types of timing…we’ll start with the holiday one*.

10. How do you think this type of visit of going to the houses of people with TB to check others in the house for TB on the holidays will be accepted by this community?

Probe: Would people in this community be willing to have someone come to their household during the holidays to check for TB, and why or why not?

Probe: What concerns might people in this community have about health workers going to the houses of people with TB to check others in the house for TB on the holidays?

11. What are the things that people in this community might like about this type of visit?

Probe: What are the benefits?

12. What are some of the things that people in this community might not like about this type of visit to check people in the household for TB?

Probe: What are the things that might not be good about this type of visit during the holidays?

*Next let’s talk about the off-peak timing [remind of definition]*.

13. How do you think going to the houses of people with TB to check others in the house for TB on the evenings and weekends will be accepted by this community?

Probe: Would people in this community be willing to have someone come to their household evenings and weekends to check for TB, and why or why not?

Probe: What concerns might people in this community have about a health worker going to the houses of people with TB to check others in the house for TB on evenings and weekends?

14. What are the things that people in this community might like about this type of visit to check people in the household for TB?

Probe: What are the benefits?

15. What are some of the things that people in this community might not like about this type of visit to check people in the household for TB?

Probe: What are the things that might not be good about this type of visit during evenings or weekends?

*Moving on to the routine type of visit [remind of definition]*.

16. How do you think the routine type of visit is accepted by this community?

Probe: Are people in this community be willing to have someone come to their household during normal business hours to check for TB, and why or why not?

Probe: What concerns do you think people in this community have about this type of visit to check people in the household for TB?

17. Thinking about all three types of timing for household visits – holiday, off-peak, and routine – which approach would you choose as the best for your community, and why?

Probe: Tell me about your choice…what would make that the best type of timing over the others?

> *Thank you all for your time…this information has been very valuable.*

